# Improvised AChE assay on homogenized rectal biopsy: An Alibi to AChE histochemistry in the diagnosis of Hirschsprung’s disease?

**DOI:** 10.1101/2023.05.11.23287813

**Authors:** Neha Kuvelkar, Serena D Souza, Maria Frances Bukelo, Vinay Jadhav, Narendra Babu, Gowri Shankar, Santosh Noronha, Usha Kini

## Abstract

The surgical treatment of Hirschsprung’s disease (HD) is reliant on the accurate tissue diagnosis based on increased acetylcholine esterase enzyme (AChE) activity in frozen rectal biopsy sections demonstrated by histochemical staining techniques. The latter techniques used for ascertaining the presence of AChE mandates the use of inaccessible sophisticated instruments thereby limiting the performance of this technique to urban resource rich settings. This bottleneck translates into delayed treatment for patients who live in suburban, resource limited regions. Given the indispensable nature of the surgical treatment, scope for improving accessibility is restricted to the diagnostic modality of the disease.

In our study, we attempted to quantitatively estimate AChE in homogenized rectal mucosal biopsy samples of paediatric population (n=68) using an in-house modified AChE (mACHE) assay. The assay procedure for homogenization of the tissue was modified to selectively extract the mucosal portion of the rectal mucosal biopsy sample to be used for the assay. The efficiency of specific mucosal extraction was confirmed by the absence of neurotrophic AChE nerve fibres in remnant tissue obtained post homogenization by histochemical AChE staining. The mAChE assay was carried out in a blinded manner and the results were compared with the gold standard histochemical AChE (hAChE) assay.

Samples with activity lesser than 1 × 10^-6^ mole/min/g wet weight of tissue were diagnosed as Non Hirschsprung’s Disease (NHD) and those above 1 × 10^-6^ mole/min/g wet weight of tissue were diagnosed as HD. mAChE assay was found to have a specificity, sensitivity of 81.08 % and 92.85%; positive, negative predictive values of 78.78% and 93.75 % respectively. The mAChE assay was able to diagnose three cases which were deemed inconclusive as per hAChE.

The improvised in-house mAChE assay had a mutually exclusive range to differentiate between control and test, thus crediting its use as a diagnostic tool for Hirschsprung’s Disease.

## Introduction

Hirschsprung’s disease (HD), characterized by the absence of myenteric and submucosal ganglionic cells involves various length of the bowel starting from distal rectum causing enterospasm and thus leading to obstruction of stool passage resulting in symptoms such as constipation, abdominal distension and vomiting associated with growth failure. It is known to affect 1 in 5000 live births and is initially suspected on failure of the new born to pass meconium (the first stool) along with abdominal distension. This condition mandates timely surgical intervention. If left undiagnosed and untreated, the disease can lead to mortality due to enterocolitis, bowel perforation and peritonitis (Heuckroth, 2018).

Diagnosis of HD is crucial and results from the combined effort of surgeons and histopathologists. This process involves evaluating the rectal mucosal biopsy from the distal rectum by an experienced histopathologist. Frozen sections cut from the well oriented rectal mucosal biopsies which are in the range of 0.2 to 0.4 cm using a cryostat, requires skill and expertise. These biopsy sections subsequently are stained with rapid Haematoxylin – Eosin stain for adequacy. Once confirmed, cut frozen sections are subjected thereafter, for rapid modified Acetylcholinesterase (AChE) histochemistry. This AChE staining technique which takes approximately 40 minutes has been improvised for use in a general pathology lab using the method of Kini *et al*.,2010, for evaluating increase in AChE activity in biopsies of HD for the final tissue diagnosis. The outcome of these two staining procedures, as ascertained by the trained histopathologists, is crucial in confirming the disease which is the gold standard for diagnosis of HD. This diagnosis is imperative as it dictates the type of management to ameliorate the symptoms i.e. medical (conservative) for non-HD cases vs surgical resection of the affected aganglionic segment (pull-through surgery), the only treatment for HD in the present scenario. Most often the two centres namely, the surgical centre where the patient and surgeon are and the other where the diagnostic lab facility is, are located at different geographical locations. The entire procedure in an Indian setting can take up from a minimum of an hour to a week or 10 days depending on the location of the patient as the diagnostic facility is not available in all centres.

While the nature of the treatment cannot be changed, we envisage that the diagnostic procedure could be improved upon. The bottleneck in the current diagnostic system is the availability and accessibility of the instrumentation required for sectioning of the frozen rectal biopsy tissue and the expertise required to have enzyme histochemistry in a general pathology lab. This is evident by the large number of cases which get directed to this tertiary referral centre which is the only diagnostic centre for gut motility disorders in the country. Additionally, the current diagnostic protocol is heavily dependent on the enzyme histochemistry-based-approach for AChE requiring the availability of trained histopathologists or calretinin based diagnostic protocol on rectal biopsy requiring a minimum of three to four days, which is an impediment curtailing diagnosis for many a child where these facilities are not available. These constraints in the existing diagnostic tools prompted us to extrapolate assessment of AChE with a different perspective and aimed to device a minimally invasive (blood-based) technique that is interpretable by a clinician. A mutually exclusive AChE activity range in plasma and red blood cells (RBC) samples of normal Indian children without gut motility/liver/hematological disorders was concluded and the simple modified improvised rapid assay protocol which was economically viable (INR 10) was devised (Kuvelkar *et al*., 2021). However, the AChE enzyme levels in plasma and in RBC, obtained in patients with HD in this study, were not statistically significant to employ it as a screening/diagnostic test for HD but the hunt continued on tissue biopsies.

The literature search touches upon the efforts made to homogenize the rectal biopsy tissue and quantitatively estimate AChE levels biochemically in occasional reports on small sample size (Boston *et al*., 1974; Dale *et al*., 1977 and 1979) but they have neither been validated nor translational. This present study has aimed to assess the best use of the developed AChE assay for screening HD patients thus simplifying the diagnostic protocol for HD in children.

## Material and Methods

The prospective double-blind study was executed in the tertiary referral health care centre after obtaining Institutional Ethical approval (IEC Study Ref. No.23/2017).

Neonates and infants who were referred to this tertiary children’s hospital for subacute intestinal obstruction presenting with symptoms such as chronic constipation, abdominal distension and vomiting and not responding to medical line of management were recruited for the study after informed consent.

Those children having one or more of the following and suspected of HD were included in the study:

1. Failure to pass meconium within 24 hrs of birth
2. Abdominal distension/vomiting, mostly non-bilious and constipation dating back to early infancy
3. Symptoms mentioned above with associated dysmorphology, trisomy 21 and stigmata of neurocristopathies or syndromic
4. Child with constipation and having a sibling with total colonic aganglionosis or long segment HD.

Those rectal tissue samples that were not received or not maintained in cold chain during transport to the lab or inadequate (lacking mucosal component) due to either ulceration or denudation resulting from frequent bowel washes were excluded from the study.

The transanal rectal mucosal biopsy samples received at the translational centre were received wrapped in saline-soaked gauze in cold chain. One of the two biopsy samples received was processed immediately for diagnostic purpose using Acetylcholine enzyme histochemistry method of Kini *et al*., 2010 and the other biopsy was stored at – 80 ^°^C for further workup of AChE biochemical assay in this study. The algorithmic approach to the tissue sample described above is depicted in Figure 1a and b.

**Figure 1A:**
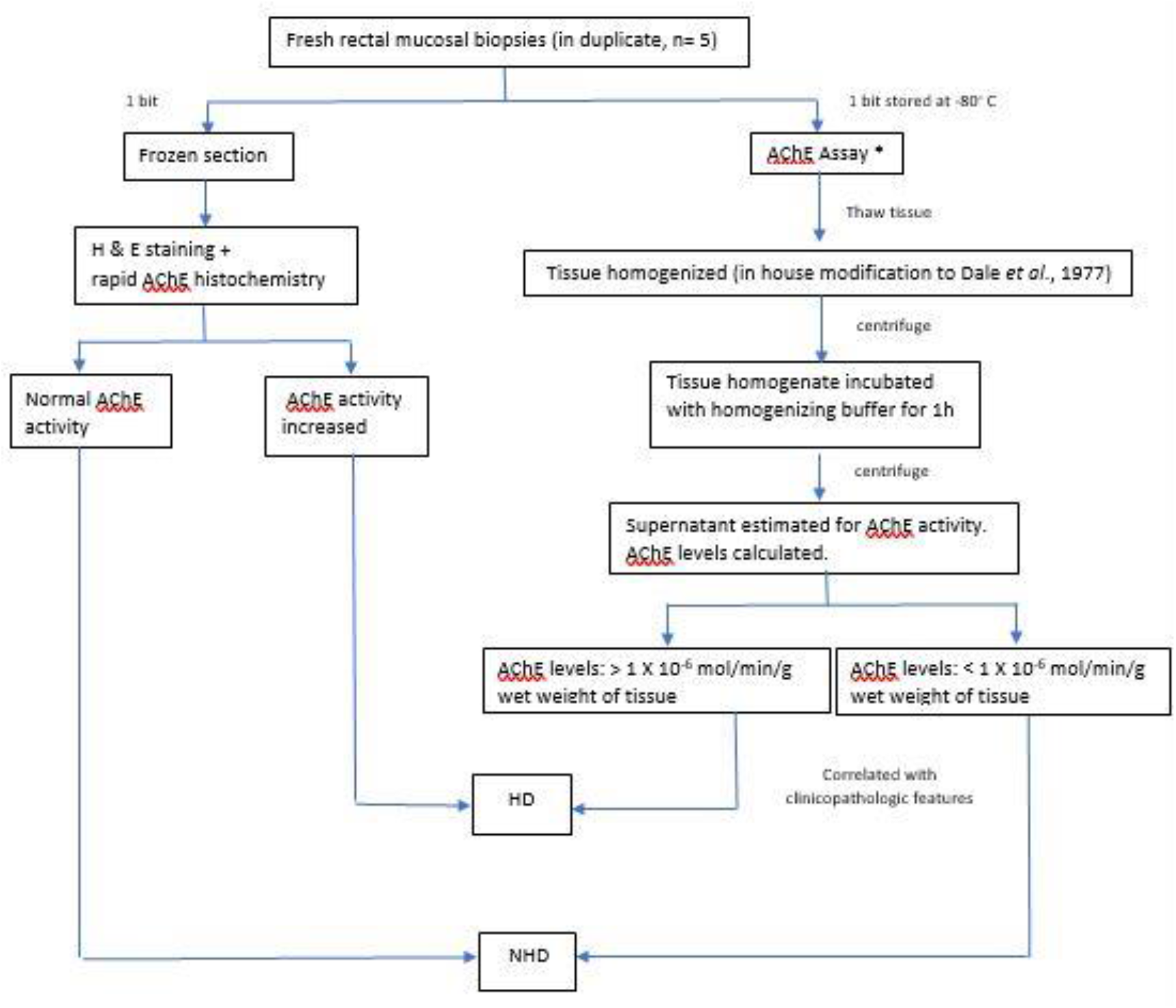

**Figure 1B:**
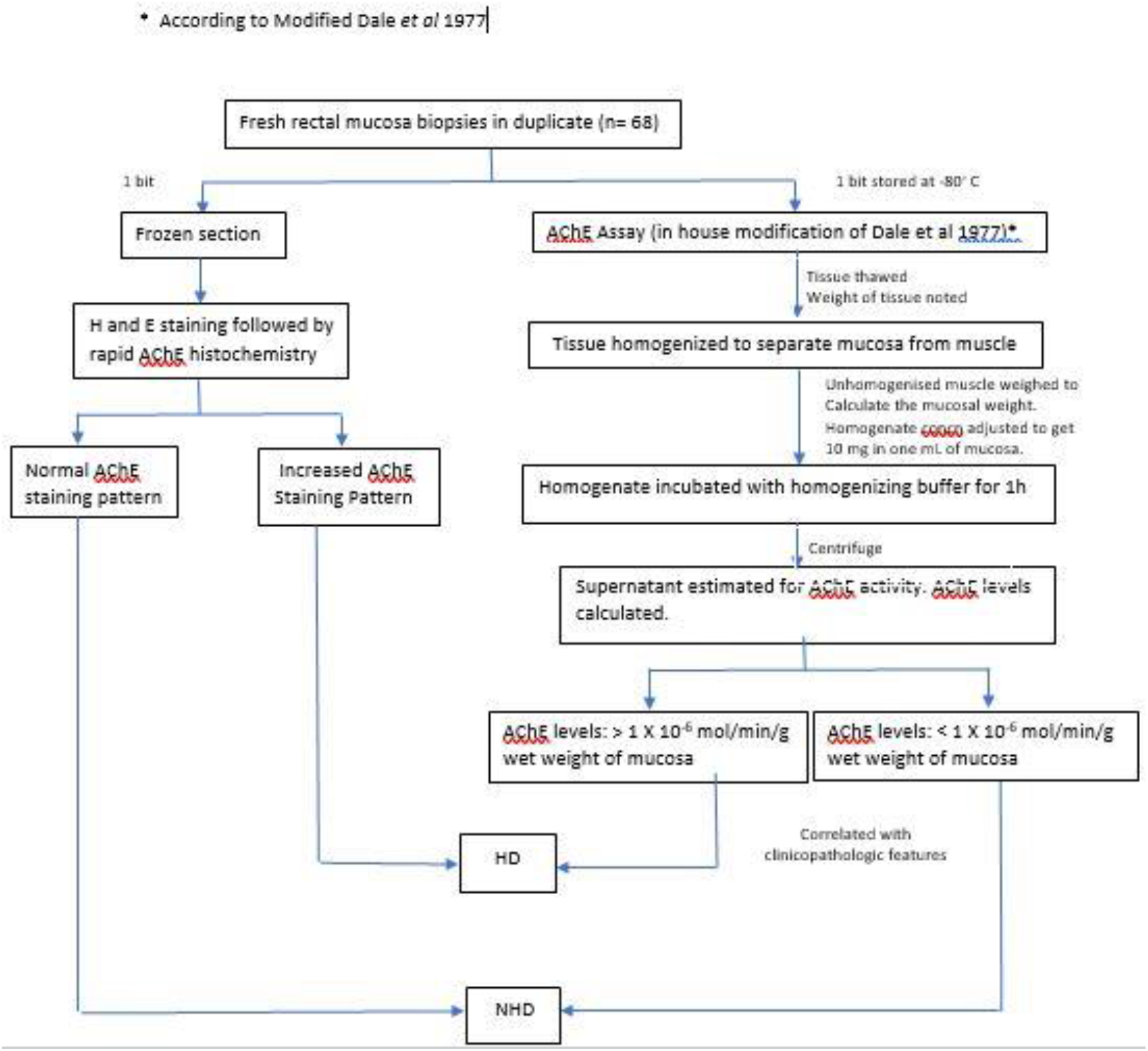

### AChE Histochemistry on frozen sections (hAChE) on transanal rectal mucosal biopsies for diagnostic purpose

Assessment for AChE activity for diagnostic purposes was carried out as follows:

Frozen sections were cut on properly oriented fresh rectal biopsies embedded in freezing medium at 10 μm thickness using Leica CM 1950 cryostat at different levels. Four even numbered sections representative of the whole biopsy were stained with rapid hematoxylin and eosin (H & E) while four odd numbered sections were stained for AChE histochemistry using rapid, improvised and modified protocols well standardized in our laboratory for final diagnosis of HD. [Babu *et al*., 2003, Yadav *et al*., 2014, Agrawal *et al*., 2015] along with a positive control slide. A frozen section from a proved case of HD was run as a positive control with every batch of sections stained for AChE which showed increased AChE activity, seen as presence of minute black fibres in the muscularis mucosa and lamina propria while the absence of the black fibres in the muscularis mucosa and lamina propria was interpreted as NHD. The histopathology report was conveyed to the surgeon for further management of the child. The frozen tissue remains/ sections were preserved at -20°C for further use.

However, the histopathology reports were kept blinded till the tissue AChE assays were completed and the values obtained were then correlated.

### AChE assay on homogenised mucosal rectal biopsies: – Phase 1

The AChE assay on the rectal tissue was performed with reference to Dale *et al*., 1977 with modifications.

The modifications (Table 1) included reduction of total assay volume, use of homogenizing buffer with Triton X – 100 as opposed to Phosphate buffer, homogenization speed and time, centrifugation time at which the assay was performed. The assay was carried out in a 3 mL cuvette which mandated the use of higher volume of tissue extract (0.4 mL) and proportionately other reagents.

**Table 1:** Modifications made in the Dale *et al*., 1977.

|  | **Phase 1 (n=5)** | Dale *et al.*, 1977 | In-house modification |
| --- | --- | --- | --- |
| 1. | Assay volume (μL) | 3000 | 300 |
| 2. | Homogenizing buffer | 0.1 M phosphate buffer (pH 8) | 0.01M Tris HCl, 1M NaCl, 0.01M EDTA and 1% Triton X-100 in distilled water (pH 7.4) |
| 3. | Centrifugation speed and time<br>For homogenization<br>For supernatant extraction | 2300 rpm X 3 '<br>14000 rpm X 2 ' | 4000 rpm X 3 – 5 'with a 15- 30 s pulse on ice<br>14,000 rpm X 15' |
| 4. | Temperature | 25 | 37 |
|  | **Phase 2 (n=68) \*** |  |  |
| 1. | Weight considered for the assay | Whole weight of tissue | Mucosal weight of tissue |
| 2. | Initial volume of homogenization buffer added for homogenization | Total volume as per the weight of the tissue | 100 μL - when tissue weighs less than 20 mg<br>200 μL - when tissue weighs more than 20 mg |

\*In addition to points 1- 4 of phase 1

Given our paediatric population of study which involved neonates, it was imperative to reduce the reaction volume to enable its use in microwell ELISA plates. Tris EDTA based buffer was used as opposed to Phosphate buffer as Tris based buffers enabled easier lysis of cells thereby enabling better extraction of the enzyme. Pulse homogenization of the tissue biopsy on ice at higher speed (4000 rpm vs 1500 rpm) was carried out to ensure efficient homogenization of the tissue. The centrifugation time was increased to obtain tissue extract supernatant without the presence of micro tissue debris. The temperature at which the reaction was carried out was raised to 37 degrees as opposed to 25 degrees. This change of temperature was adopted keeping in mind that the hAChE histochemical stain is also carried out at 37°C to stain for AChE (Kini *et al*., 2010).

With the above improvisations and modifications incorporated, the methodology employed was as follows: The rectal biopsy tissue was thawed on ice, blot dried on tissue paper and weighed. It was then transferred into a homogenising cup kept on ice to which 1 mL of homogenising buffer (0.01M Tris HCl, 1M NaCl, 0.01M EDTA and 1% Triton X-100 in distilled water, pH 7.4) per 10 mg of tissue was added. The tissue was homogenized at 4000 rpm till complete homogenization was achieved using RQT-127A homogeniser (Remi Lab World, India). The homogenate was centrifuged at 14,000 rpm for 15 minutes at 4 °C in a cold centrifuge (Thermo Fisher Scientific, Roskilde, Denmark). The obtained supernatant was estimated for tissue AChE enzyme activity at 37°C by measuring absorbance change per minute for 5 minutes at 412 nm using the procedure as described in Table 2. Further quantification was carried out using the formula.

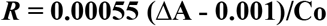

where ***R =*** mole substrate hydrolysed /min/ g wet weight of tissue,

Co = observed tissue concentration in mg/mL,

ΔA = observed absorbance change per minute.

Rate of non-enzymic hydrolysis under the conditions of the assay to be 0.001 ***A*** units/min.

The results were reported in terms of AChE activity, R × 10^-6^

**Table 2:**
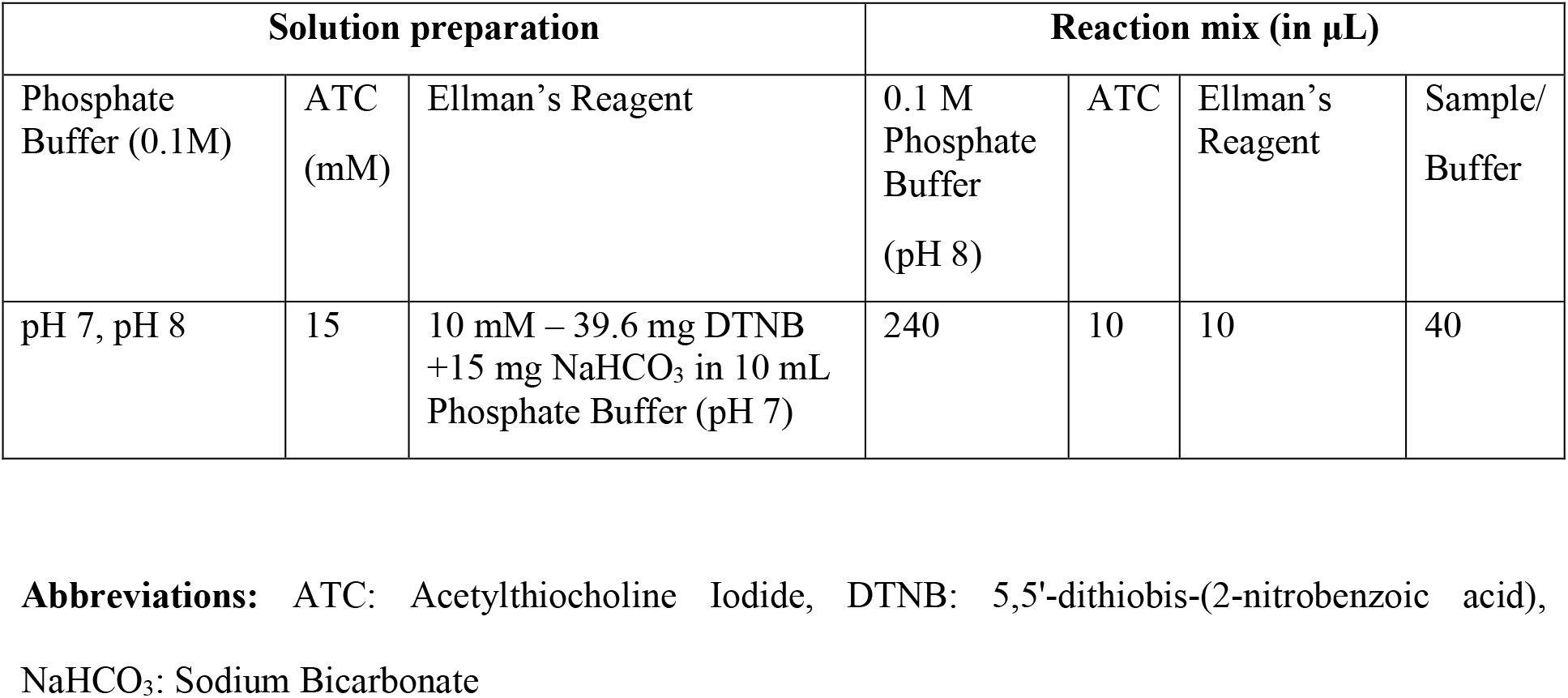
mAChE enzyme assay.

| Solution preparation | | | Reaction mix (in $\mu\text{L}$) | | | |
| --- | --- | --- | --- | --- | --- | --- |
| Phosphate Buffer (0.1M) | ATC (mM) | Ellman's Reagent | 0.1 M Phosphate Buffer (pH 8) | ATC | Ellman's Reagent | Sample/ Buffer |
| pH 7, pH 8 | 15 | 10 mM – 39.6 mg DTNB +15 mg $\text{NaHCO}_3$ in 10 mL Phosphate Buffer (pH 7) | 240 | 10 | 10 | 40 |

**Abbreviations:** ATC: Acetylthiocholine Iodide, DTNB: 5,5'-dithiobis-(2-nitrobenzoic acid), $\text{NaHCO}_3$ : Sodium Bicarbonate

Post the assay, the samples were classified as HD /NHD based on a classifier range of 1 × 10^-6^ mole/min/g wet weight of tissue (Dale *et al*., 1977). Samples with activity lesser than 1 × 10^-6^ mole/min/g wet weight of tissue were diagnosed as NHD and those above 1 × 10^-6^ mole/min/g wet weight of tissue were diagnosed as HD. These results were then compared to the hAChE results (gold standard).

### Modified AChE assay for estimation of AChE activity in homogenized mucosal rectal biopsy (mAChE) – Phase 2

We were unable to obtain results concordant with the histopathology diagnosis when we estimated AChE activity (Phase 1) using modified Dale *et al*., 1977. This could likely to be due to the use of whole rectal biopsy tissue as opposed to the mucosal tissue wherein the enzyme predominantly resides and is easily retrievable than from muscle (muscularis propria) Also, in certain cases, the tissue samples received were very small in size and had lower weight. Adding 1mL of homogenizing buffer per 10 mg tissue might have caused the tissue to be incompletely homogenized due to increased volume of buffer in proportion to the weight of tissue.

We, therefore, modified the initial volume of buffer to be added while homogenizing based on the mucosal weight. This led to modification of the procedure further. The modifications included utilizing only the mucosa for AChE assay and the volume of homogenizing buffer initially added for the purpose of homogenization (Table 1). To elaborate the steps, the tissue was thawed on ice, blot dried on tissue paper and weighed (W1 mg). It was quickly transferred into a homogenizing cup containing homogenizing buffer (0.01M Tris HCl, 1M NaCl, 0.01M EDTA and 1% Triton X-100 in distilled water, pH 7.4). The initial volume of homogenizing buffer to be added was determined based on the initial weight (W1 mg) of the tissue. 100 / 200 μL homogenizing buffer was added when the tissue weighed less than/ more than 20 mg respectively. The tissue was homogenized at 4000 rpm with a 15-20 sec pulse while keeping the homogenizing cup on ice bath to separate mucosa from the muscle layer. The unhomogenized tissue was blot dried and weighed (W2 mg). The weight of the mucosa was obtained [W1-W2=W3] and the concentration of the obtained tissue homogenate was adjusted to 10 mg per mL with the homogenizing buffer and incubated on ice for 1 hour. The homogenate was centrifuged for 15 minutes at 14,000 rpm at 4 °C in a cold centrifuge (Thermo Fisher Scientific, Roskilde, Denmark). The supernatant, thus, obtained was further used for estimating the AChE activity at 37°C by measuring absorbance change per minute for 5 minutes at 412 nm using the procedure as described in Table 2. The AChE activity was calculated and diagnosis based considering the threshold of 1 × 10^-6^ mole/min/g wet weight of mucosa as described earlier.

The AChE enzyme activity (mol/min/g of wet weight of mucosa) was measured as absorbance change at 412 nm at 37°C over a period of 5 minutes with a 1-minute interval. Further quantification was carried out using the formula

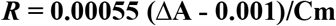

where ***R =*** mole substrate hydrolysed /min/ g wet weight of mucosa, Cm = observed mucosal concentration in mg/mL,

ΔA = observed absorbance change per minute.

Rate of non-enzymic hydrolysis under the conditions of the assay to be 0.001 ***A*** units/min.

The results were reported in terms of AChE activity, R × 10^-6^ mole/min/g wet weight of mucosa

### Remnant tissue analysis

After performing the homogenization as per Phase 2, the remnant tissue wherever available (for few randomly selected) were studied histopathologically for morphology (Haematoxylin and Eosin staining) and its AChE activity by freezing the tissue and staining the frozen sections in similar lines to the diagnostic biopsies. This was done to confirm the constituents of the remnant tissue namely smooth muscle bundles and that mucosa was completely stripped and homogenised and therefore separated from the tissue thereby confirming that the AChE estimation was exclusively from the mucosa and not from the muscle layer (muscularis propria) (Table 4).

### Performance parameters of the in-house modified Tissue AChE assay

Sensitivity, specificity and prediction of the tissue AChE assay were as follows:

**Sensitivity**= Number of correctly predicted HD cases /Number of True HD cases*100

**Specificity**= Number of correctly predicted NHD cases / Number of True NHD cases*100

**Prediction accuracy**= Number of correctly predicted cases/Number of total cases *100

**Positive Predictive Value (PPV)** = Number of true positives cases /Total positive cases * 100

**Negative Predictive Value (NPV)** = Number of true negatives cases / Total negative cases * 100

## RESULTS

### Tissue AChE histochemistry (hAChE)

AChE histochemistry (hAChE) on rectal biopsies from five and 68 cases for diagnostic purposes were read, based on the following criteria:

A rectal mucosal biopsy was considered as HD if

1. Hypertrophic/hyperplastic nerve bundles were identified in the submucosa with no ganglion cells
2. Increased AChE activity with positive staining of hypertrophic/hyperplastic nerve fibers as dark green-black staining in specific patterns of either
  i. Pattern A where nerve fibers in the submucosa extending through the muscularis mucosa into the lamina propria or
  ii. Pattern B where nerve fibers extending only up to the base of the crypts
  iii. Equivocal pattern where hypertrophic nerve bundles in the submucosa alone with no specific pattern in the lamina propria.

A rectal mucosal biopsy was considered as non-HD if

1. At least one ganglion cell was identified in one/both the H and E sections
2. The AChE stained an occasional nerve twig in the submucosa and highlighted a ganglion cell if included in the biopsy. However, no stainable AChE fibers were identifiable (negative staining) in the muscularis mucosa and lamina propria.

Two and 28 cases from Phase I & II showed increased AChE activity in the submucosa and/or mucosa with no ganglion cells respectively and were diagnosed as HD mandating surgical intervention while three and 37 of phase I & II were ganglionic and showed no increase in AChE activity respectively, thus, diagnosing them as non-HD cases. The three cases 11, 43 and 49 showed equivocal results with no pattern with AChE histochemistry. They were followed up with repeat rectal biopsies confirming them to be HD. However, we were not able to procure a corresponding rectal mucosal biopsy sample for mAChE assay and hence these samples were not included in the performance parameter calculation for the mAChE assay.

The results of the AChE tissue assay of this prospective double-blind study carried out on tissue in two phases are as follows: The first phase referred to as Phase 1 which involved using the entire homogenized rectal mucosal biopsy for AChE activity estimation. Five rectal mucosal biopsy samples received were in the age range of 0-5 years. The rectal mucosal biopsy samples used in Phase 2 wherein, only the homogenized mucosa (with no muscle) was used for AChE activity estimation were obtained from children with the mean age group of 0-5 years.

### Phase 1

The AChE activity in the tissue samples was estimated as per Dale *et al*., 1977 protocol as described in Table 1. The AChE activity results obtained from the tissue AChE assay with the gold standard are elaborated in Table 2 wherein we found the results of 2 out of 5 samples to be discordant. (Table 3).

**Table 3:** Tissue AChE assay and concordance with histopathology results.

| Sl.No. | Pattern AChE Histochemistry (HP) | Diagnosis (HP) | AChE levels (mole/min/g of tissue) X 10<sup>-6</sup> | Diagnosis (Assay) | Discordance |
| --- | --- | --- | --- | --- | --- |
| Phase 1 |  |  |  |  |  |
| 1 | A | HD | 1.64 ± 0.007 | HD | No |
| 2 | Negative E | NHD | 0.66 ± 0.01 | NHD | No |
| 3 | Negative E | NHD | 1.56 ± 0.03 | HD | Yes |
| 4 | E | HD | 0.25 ± 0.03 | NHD | Yes |
| 5 | Negative E | NHD | 0.68 ± 0.07 | NHD | No |
| Phase 2 |  |  |  |  |  |
| 1 | B to A | HD | 3.47 ± 0.01 | HD | No |
| 2 | B to A | HD | 24.44 ± 0.98 | HD | No |
| 3 | A | HD | 3.36 ± 0.05 | HD | No |
| 4 | A | HD | 5.94 ± 0.07 | HD | No |
| 5 | Negative E | NHD | 2.26 ± 0.08 | HD | Yes |
| 6 | Negative E | NHD | 0.14 ± 0.04 | NHD | No |
| 7 | Negative E | NHD | 0.23 ± 0.05 | NHD | No |
| 8 | Negative E | NHD | 0.34 ± 0 | NHD | No |
| 9 | Negative E | NHD | 0.34 ± 0.04 | NHD | No |
| 10 | Negative E | NHD | 0.26 ± 0.01 | NHD | No |
| 11 | Neither ganglion cells nor increase in AChE | Inconclusive | 1.64 ± 0.16 | HD | - |
| 12 | A | HD | 0.13 ± 0.007 | NHD | Yes |
| 13 | Negative E | NHD | 0.52 ± 0.07 | NHD | No |
| 14 | Negative E | NHD | 0.19 ± 0.16 | NHD | No |
| 15 | Negative E | NHD | 0.62 ± 0 | NHD | No |
| 16 | A | HD | 6.99 ± 0.07 | HD | No |
| 17 | B | HD | 2.18 ± 0.07 | HD | No |
| 18 | Negative E | NHD | 0.73 ± 0.05 | NHD | No |
| 19 | E | HD | 9.33 ± 0.11 | HD | No |
| 20 | Negative E | NHD | $0.95 \pm 0.04$ | NHD | No |
| 21 | Negative E | NHD | $5.37 \pm 0.03$ | HD | yes |
| 22 | Negative E | NHD | $0.17 \pm 0.01$ | NHD | No |
| 23 | B to A | HD | $7.32 \pm 0.05$ | HD | No |
| 24 | A | HD | $6.78 \pm 0.03$ | HD | No |
| 25 | Negative E | NHD | $0.34 \pm 0.09$ | NHD | No |
| 26 | A | HD | $4.57 \pm 0.02$ | HD | No |
| 27 | B to A | HD | $2.86 \pm 0.1$ | HD | No |
| 28 | Negative E | NHD | $2.18 \pm 0.03$ | HD | Yes |
| 29 | E | NHD | $0.73 \pm 0.15$ | NHD | No |
| 30 | Negative E | NHD | $0.27 \pm 0.007$ | NHD | No |
| 31 | Negative E | NHD | $1.56 \pm 0.07$ | HD | Yes |
| 32 | A | HD | $1.11 \pm 0.09$ | HD | No |
| 33 | A | HD | $0.63 \pm 0.04$ | NHD | Yes |
| 34 | A | HD | $1.01 \pm 0.06$ | HD | No |
| 35 | Negative E | NHD | $0.63 \pm 0.05$ | NHD | No |
| 36 | A | HD | $1.02 \pm 0.01$ | HD | No |
| 37 | A | HD | $9.60 \pm 4.29$ | HD | No |
| 38 | Negative E | NHD | $0.73 \pm 0.01$ | NHD | No |
| 39 | Negative E | NHD | $0.63 \pm 0.04$ | NHD | No |
| 40 | A | HD | $4 \pm 0.07$ | HD | No |
| 41 | Negative E | NHD | $0.67 \pm 0$ | NHD | No |
| 42 | Negative E | NHD | $0.52 \pm 0.01$ | NHD | No |
| 43 | No pattern | Inconclusive | $2.45 \pm 0.18$ | HD | - |
| 44 | Negative E | NHD | $0.73 \pm 0.007$ | NHD | No |
| 45 | A | HD | $2.7 \pm 0.09$ | HD | No |
| 46 | Negative E | NHD | $0.86 \pm 0.26$ | NHD | No |
| 47 | Negative E | NHD | $0.71 \pm 0.1$ | NHD | No |
| 48 | A | HD | $2.51 \pm 0.15$ | HD | No |
| 49 | No pattern | Inconclusive | $2.83 \pm 0.31$ | HD | - |
| 50 | Negative E | NHD | $1.60 \pm 0.01$ | HD | Yes |
| 51 | Negative E | NHD | $1.24 \pm 0.06$ | HD | Yes |
| 52 | Negative E | NHD | $0.55 \pm 0.06$ | NHD | No |
| 53 | Negative E | NHD | $0.92 \pm 0.007$ | NHD | No |
| 54 | B to A | HD | $2.98 \pm 0.21$ | HD | No |
| 55 | E | HD | $2.12 \pm 0.59$ | HD | No |
| 56 | A | HD | $9.02 \pm 1.54$ | HD | No |
| 57 | Negative E | NHD | $0.51 \pm 0.05$ | NHD | No |
| 58 | Negative E | NHD | $0.61 \pm 0.03$ | NHD | No |
| 59 | Negative E | NHD | $0.37 \pm 0.07$ | NHD | No |
| 60 | Negative E | NHD | $1.32 \pm 0.23$ | HD | Yes |
| 61 | Negative E | NHD | $0.29 \pm 0.08$ | NHD | No |
| 62 | Negative E | NHD | $0.43 \pm 0.007$ | NHD | No |
| 63 | B | HD | $1.66 \pm 0.09$ | HD | No |
| 64 | E | HD | $1.56 \pm 0.22$ | HD | No |
| 65 | A | HD | $2.36 \pm 0.07$ | HD | No |
| 66 | Negative E | NHD | $0.61 \pm 0.01$ | NHD | No |
| 67 | A | HD | $3.52 \pm 0.04$ | HD | No |
| 68 | A | HD | $3.52 \pm 0.4$ | HD | No |

**Table 4:** Morphology and AChE enzyme staining performed on remnant tissue samples post homogenization.

| **Sl.no.** | **Haematoxylin and Eosin staining of the remnant tissue** | **AChE staining of the remnant tissue** |
| --- | --- | --- |
| 31 | Muscle + scanty submucosa +<br>Mucosa - | Negative |
| 32 | Muscle + scanty submucosa+<br>Mucosa - | Negative |
| 33 | Mucosa -<br>Muscle +, scanty submucosa + | Focal increase in AChE<br>positive |
| 35 | Submucosa minute fragment,<br>mucosa - | Negative |
| 36 | Scanty Submucosa +, mucosa - | Negative |
| 37 | Muscle + scanty submucosa - | Negative |
| 38 | Muscle + scanty submucosa+<br>Mucosa - | Negative |
| 39 | Neither submucosa nor scanty muscle<br>could be identified, mucosa - | Negative |

+ present
- absent

### Phase 2

The modified assay procedure referred to as mAChE assay was used to assess the AChE activity of 68 rectal mucosal biopsy samples and the obtained AChE activity subjected to a classifier of 1 × 10^-6^ mol/min/mg of mucosal weight, showed the following results; 36 samples were found to have AChE activity above the classifier with a mean of 3.42 × 10 ^-6^ ± 2.43 10 ^-6^ mol/min/g of wet weight of mucosa (with an outlier of 24.44 × 10 ^-6^ mol/min/g wet weight of mucosa) and were therefore concluded to be HD on the AChE values whereas 32 samples were found to have activity below the classifier with a mean of 0.48 × 10 ^-6^ ± 0.24 × 10 ^-6^ mol/min/g of wet weight of mucosa and were deemed to be NHD (Table 3). On comparing with the gold standard, we observed that of the 36 suspected HD cases as per mAChE assay, 26 were correctly predicted as HD and 7 cases were falsely predicted as HD (false positives).. However, the mAChE assay indicated the three inconclusive biopsies (proved as HD in the subsequent second biopsy to be HD) as HD on the biopsy without repeats. Of the 32 supposed NHD cases as per mAChE assay, 30 cases were correctly predicted as NHD and 2 cases were incorrectly predicted as NHD (false negatives). The correlated mAChE activity levels with the histochemistry is depicted in Table 3. The patterns obtained from hAChE and the AChE values obtained from mAChE assay have been plotted (Figure 2).

**Figure 2:**
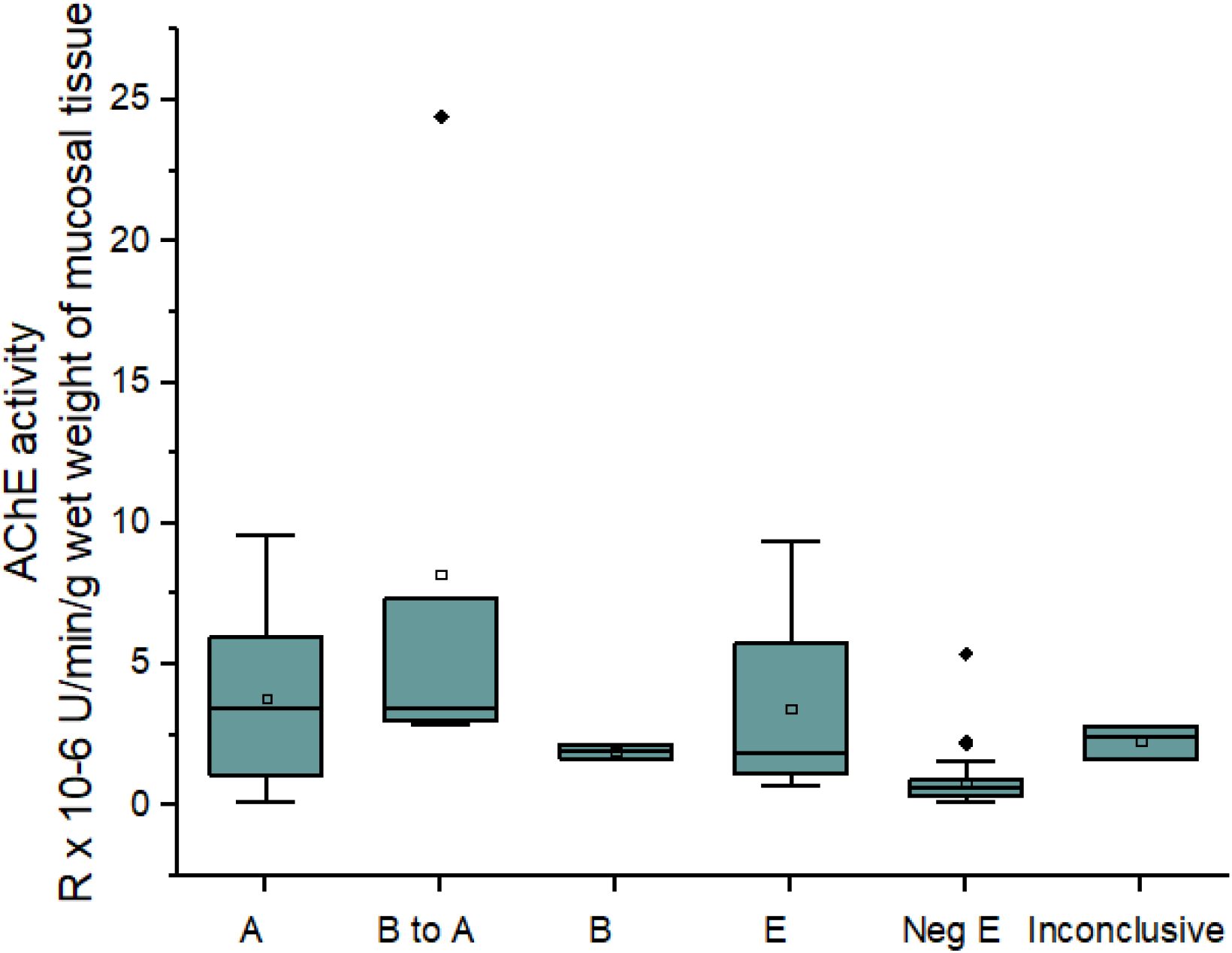
AChE pattern and AChE assay levels as obtained by hAChE and mACHE respectively.

We calculated the performance parameters of the mAChE assay and found the mAChE assay to have a specificity of 81.08 % and sensitivity of 92.85% (Table 5); positive predictive value of 78.78 % and negative predictive value of 93.75 %.

**Table 5:** Performance characteristics of mAChE assay.

|  | No of samples |  |  |
| --- | --- | --- | --- |
|  | HD | NHD | Inconclusive |
| **Histopathology (n=68)** | 28 | 37 | 3 |
| **Tissue AChE Assay (n=68)** | 36 | 32 | - |
| **Tissue AChE Assay (n=65)\*** |  |  |  |
| **True positives** | 26 |  |  |
| **True negatives** | 30 |  |  |
| **False positives** | 7 |  |  |
| **False negatives** | 2 |  |  |
| **Sensitivity** | 92.85 % |  |  |
| **Specificity** | 81.08 % |  |  |
| **Positive Predictive Value (PPV)** | 78.78% |  |  |
| **Negative Predictive Value (NPV)** | 93.75 % |  |  |
| **Prediction accuracy** | 86.15% |  |  |

\*3 inconclusive cases not included in the statistics
HD: Hirschsprung's Disease; NHD: Non- Hirschsprung's Disease

## Discussion

The diagnosis of Hirschsprung’s disease is confirmed by staining for an enzyme, acetylcholine esterase, on frozen sections of rectal mucosal biopsies. Hypertrophic AChE nerve bundles in the lamina propria and muscularis mucosa have been observed in rectal mucosal biopsy of patients suffering from Hirschsprung’s Disease (Dale *et al*., 1979; Babu *et al*.,2003). This technique requires the availability of high-end cryostat for sectioning the tissue which may be a major bottleneck in most primary healthcare centres. Given that the AChE enzyme is localized in mucosa of the rectal biopsy, the AChE enzyme can be studied by performing an enzyme assay in the homogenized tissue as suggested by Dale *et al*., 1977 with modifications. On performing the AChE assay, we observed incorrect diagnosis of Hirschsprung’s Disease in 2 of 5 samples when the whole rectal mucosal biopsy tissue was homogenized and used for AChE activity determination as specified in the protocol by Dale *et al*., 1977. We attributed this error to two parameters of the assay. The first parameter was the initial volume of homogenization buffer to be added while homogenizing the tissue. We introduced a classifier based on the weight of the mucosal tissue. Homogenizing buffer (100/200 μL) was added when the tissue weighed below or above 20 mg of mucosa respectively. The second parameter was the part of the homogenized tissue to be used for AChE activity estimation. We observed the mucosa to homogenize much faster than the rather stubborn muscularis propria layer. Given that the hypertrophic nerve bundles are primarily localized in the mucosa (Kakita 2000; Park 1992; Barr 1985), we used only the homogenized mucosa for AChE activity estimation. The presence of the mucosal layer reported to be vital to the assay as histopathology studies have reported that insufficient mucosa in a biopsy could lead to false positive interpretations of results (Mukhopadhyay *et al*., 2017). We further confirmed the homogenization of the mucosal layer by performing a Haematoxylin and Eosin staining as well as an AChE histochemistry on the remnant muscle layer obtained post homogenization.

By implementing these modifications, the improvised AChE assay helped us to report correct diagnosis in 56 of 68 rectal mucosal biopsy samples which was a significant and satisfactory improvement (Table 3) with a mutually exclusive AChE activity level between the control and the test samples. We further analysed the 9 incorrect diagnoses (7 false positives and 2 false negatives) as per the mAChE assay. On detailed analysis, we observed that all the false positive cases had equivocal pattern E in the AChE histochemistry stain and yet had AChE levels higher than the classifying threshold.

## Conclusion

In summary, the mAChE assay improvised in-house has a mutually exclusive range to differentiate between Hirschsprung’s disease and non-Hirschsprung’s disease, thus crediting its use as a diagnostic tool for Hirschsprung’s disease. The assay was also able to overcome the challenges faced by the three in-conclusive cases diagnosed using histopathology. Though the improvised mAChE assay, cannot differentiate between AChE patterns as noted in histopathology, given it’s sensitivity of 92.85% and positive predictive value of 78.78 %, it is of profound use for timely diagnosis when histopathology services are not available at peripheral laboratories.

## Data Availability

All data produced in the present work are contained in the manuscript

## References

Agrawal RK, Kakkar N, Vasishta RK, Kumari V, Samujh R and Rao KLN. Acetylcholinesterase histochemistry (AChE) - a helpful technique in the diagnosis and in aiding the operative procedures of Hirschsprung disease. Diagn Pathol., 2015; 10:208.

Babu MK, Usha Kini, Das K, Alladi A and D’Cruz A. A modified technique for the diagnosis of Hirschsprung’s disease from rectal biopsies. National Med Jr India 2003; 16: 245–248.

Barr LC, Booth J, Filipe MI and Lawson JO. Clinical evaluation of the histochemical diagnosis of Hirschsprung’s disease. Gut, 1985; 26(4):393–399

Boston VE, Dale G and Riley KWA. Diagnosis of Hirschsprung’s disease by quantitative biochemical assay of AChE in rectal tissue. Lancet 1975; 2: 951–953.

Dale G, Bonham JR, Lowdon P, Wagget J, Rangecroft L and Scott DJ. Diagnostic value of rectal mucosal acetylcholinesterase levels in Hirschsprung’s Disease. Lancet, 1979; 347–349.

Dale G, Bonham JR, Riley KWA and Wagget J. An improved method for the determination of Acetylcholinesterase activity in rectal biopsy tissue from patients with Hirschsprung’s disease. J.Clinica Chimica Acta, 1977; 77(3):407–413.

Heuckeroth RO. Hirschsprung disease — integrating basic science and clinical medicine to improve outcomes. Nature reviews, Gastroenterology & hepatology; 2018; 15: 152–167

Kakita Y, Oshiro K, O’Briain DS and Puri P. Selective Demonstration of Mural Nerves in Ganglionic and Aganglionic Colon by Immunohistochemistry for Glucose Transporter-1 Prominent Extrinsic Nerve Pattern Staining in Hirschsprung Disease. Arch Pathol Lab Med 2000; 124:1314–1319.

Kini U, Das K, Babu MK, Mohanty S, Divya P and Saleem KM. Role of rapid modified acetylcholineesterase histochemistry in the diagnosis of Hirschsprungs disease. Indian J Path Microbiol. 2010;53:s127.

Kuvelkar N, Dsouza S, Vidhyashree K, Shankar G, Bukel MF, Noronha S and Kini U. Estimation of plasma and RBC acetylcholinesterase in children: An evaluation tool for Hirschsprung’s disease? Indian Journal of Pathology and Microbiology, 2021:64(2): 266–276.

Martucciello G, Prato AP, Puri P, Holschneider AM, Meier-Ruge W, Jasonni V, et al. Controversies concerning diagnostic guidelines for anomalies of the enteric nervous system: A report from the fourth International Symposium on Hirschsprung’s disease and related neurocristopathies. J Pediatr Surg 2005; 40:1527–31.

Mukhopadhyay, M Sengupta, C Das, M Mukhopadhyay, S Barman and B Mukhopadhyay. Immunohistochemistry-based comparative study in detection of Hirschsprung’s disease in infants in a Tertiary Care Center. Journal of Laboratory Physicians, 2017; 9(2): 76–80

Park, WH, Choi SO, Kwon KY and Chang ES. Acetylcholinesterase histochemistry of rectal suction biopsies in the diagnosis of Hirschsprung’s disease. J Korean Med Sci, 1992; 7: 353–359.

Yadav L, Kini U, Das K, Mohanty S and Puttegowda D. Calretinin immunohistochemistry versus improvised rapid Acetylcholinesterase histochemistry in the evaluation of colorectal biopsies for Hirschsprung disease. Indian Journal of Pathology and Microbiology, 2014; 57(3): 369–375

